# Nanopore Sequencing of SARS-CoV-2: Comparison of Short and Long PCR-tiling Amplicon Protocols

**DOI:** 10.1101/2021.05.12.21256693

**Authors:** Broňa Brejová, Kristína Boršová, Viktória Hodorová, Viktória Čabanová, Askar Gafurov, Dominika Fričová, Martina Neboháčová, Tomáš Vinař, Boris Klempa, Jozef Nosek

## Abstract

Surveillance of the SARS-CoV-2 variants including the quickly spreading mutants by rapid and near real-time sequencing of the viral genome provides an important tool for effective health policy decision making in the ongoing COVID-19 pandemic. Here we evaluated PCR-tiling of short (∼400-bp) and long (∼2 and ∼2.5-kb) amplicons combined with nanopore sequencing on a MinION device for analysis of the SARS-CoV-2 genome sequences. Analysis of several sequencing runs demonstrated that using the long amplicon schemes outperforms the original protocol based on the 400-bp amplicons. It also illustrated common artefacts and problems associated with this approach, such as uneven genome coverage, variable fraction of discarded sequencing reads, as well as the reads derived from the viral sub-genomic RNAs and/or human and bacterial contamination.

## Introduction

Massive spreading of severe acute respiratory syndrome coronavirus 2 (SARS-CoV-2) within the human population began in December 2019 in Wuhan, Hubei Province, China (1-3). In the following weeks, the virus has been quickly transmitted all over the globe. As of April 2021, it infected more than 147 million humans and caused over 3.1 million deaths (https://arcg.is/0fHmTX; 4). The 29,903-nt long genomic RNA sequence of the SARS-CoV-2 strain Wuhan-Hu-1 (Genbank/RefSeq acc.nos. MN908947 / NC_045512; 1) and related isolates (2,3) were determined early in 2020 and facilitated rapid development of molecular diagnostics as well as the analysis of additional isolates from other geographical regions of the world. Currently, more than 1 million SARS-CoV-2 genome sequences are available in the GISAID repository (http://www.gisaid.org), thus representing an unprecedented resource for the scientific community and public health officials. The virus sequences were determined using a range of experimental approaches based on metagenomics, sequence capture or enrichment, amplicon pools by deploying short (*e*.*g*., Illumina) or long-read (*e*.*g*., Pacific Biosciences, Oxford Nanopore Technologies) sequencing platforms. Of these, the nanopore sequencing becomes increasingly popular as in addition to sequencing of viral genomic RNA it also permits transcriptome mapping, characterization of sub-genomic RNA molecules, and identification of modified nucleotides in the viral genome (5-7). Rapid, cost-effective, and near real-time genome sequencing of the SARS-CoV-2 variants combined with epidemiological data provides an important resource not only for understanding the virus transmission, its genetic alterations and evolution, but also for making the policy decisions in combating the pandemic (8). Monitoring sequence diversification plays an essential role in continual refinement of molecular diagnostics (*e*.*g*., redesigning the primers for nucleic acid amplification techniques (9) or development of screening tools for variants of concerns (VoC) and those evading the immune response (10, 11)). This underscores the importance of genomic epidemiology, although the elucidation of direct links between particular mutation(s) and the virus spreading or clinical implications still represents a challenging task (12-22).

Nanopore sequencing of tiled PCR-generated amplicon pools represents a powerful tool for investigating viral genomes. The protocol has been developed by the Artic Network (https://artic.network/) for sequencing of Ebola, Zika, and Chikungunya genomes (23, 24). In January 2020, the original protocol was promptly adjusted for rapid sequence determination of SARS-CoV-2 RNA prepared directly from clinical samples such as nasopharyngeal or oropharyngeal swabs. Additional studies described its modifications including alternative primer schemes and different amplicon sizes or different sequencing chemistries (25-36). Its further improvements resulted in simplification of the sequencing library preparation, shortened hands-on time, and increased sample multiplexing (up to 96) that decreased the reagent costs to about £10 per sample, making this approach affordable for epidemiologic surveillance of the pandemics (36). Importantly, the rigorous comparison of nanopore sequencing with Illumina short reads technology demonstrated that in spite of relatively high error rates in individual nanopore reads, the highly accurate consensus single nucleotide variant (SNV) calling with >99% sensitivity and >99% precision can be achieved with a minimum of about 60-fold coverage (37).

In this study, we compare the performance of several PCR-tiling based protocols which were evaluated as part of our efforts to sequence isolates of SARS-CoV-2 from Slovakia collected between March 2020 and March 2021. Using the generated sequence data, we investigate the nature of common problems and artefacts associated with this approach. We compared the sequencing results obtained from the libraries containing multiplexed barcoded SARS-CoV-2 samples made of ∼400-bp, ∼2-kb, and ∼2.5-kb long overlapping amplicon pools as well as the combination of short and long amplicons. Our results show that sequencing of long amplicons clearly outperforms the original protocol based on shorter amplicons in terms of lower coverage variation and overall quality of the final sequence consensus. We also compare the performance of MinION runs with the standard (FLO-MIN106, FLO-MIN106D) and flongle (FLO-FLG001) flow cells differing by nominal pore counts, *i*.*e*. 2048 (split into four sets of 512 each) and 126, respectively.

## Results and Discussion

### Comparison of PCR-tiling amplification protocols

The PCR-tiling amplification combined with nanopore sequencing was employed for genome sequence analysis of 152 SARS-CoV-2 isolates from Slovakia (**S1 Table**). The genome sequences were obtained using primer schemes generating either ∼400-bp (Artic Network version V3, https://github.com/artic-network/artic-ncov2019), ∼2-kb (35), or ∼2.5-kb long amplicons (27). In some experiments, we loaded the same sequencing libraries to both the standard and Flongle flow cells. This allowed us to compare and evaluate the MinION runs and to analyze the problems affecting the performance of nanopore sequencing.

To compare primer sets for short and long amplicons and sequencing devices, three different batches (UKBA-2, UKBA-3 and UKBA-4 in **Table 1**) consisting of 10-12 multiplexed samples were sequenced using multiple strategies. **Fig 1** shows the comparison of the fraction of samples in a sequencing run successfully sequenced at various cut-offs measuring the total amount of sequencing normalized by the number of samples in the run. In both batches UKBA-2 and UKBA-3, 2-kb amplicons clearly outperform 400-bp amplicons. Sequencing of a mixture of longer and shorter amplicon pools provided comparable results to sequencing longer amplicons alone, perhaps because the mixture was enriched in the long amplicons. Finally, the Flongle and standard flow cells are similarly successful at comparable sequencing volumes. However, there are two disadvantages to using the Flongle flow cells. First, the Flongle cannot be washed and reused, its entire capacity is used for a single experiment. Second, since there is a large variance in the amount of data produced by a single Flongle flow cell (in our experiments, the number of active pores in Flongles ranged between 18 and 67 pores and produced between 110 and 830 Mbp - see Table 1), the capacity may be insufficient to completely recover sequences of 10 or more multiplexed samples. We consider as an important advantage that the runs using the standard flow cells can be terminated when sufficient data is collected, and thus these flow cells can be reused in further experiments after washing with nuclease containing buffer (*i*.*e*., EXP-WSH003 or EXP-WSH004). Moreover, the standard flow cells allow simultaneous sequencing of a greater number of barcoded samples with a longer run.

**Table 1.**
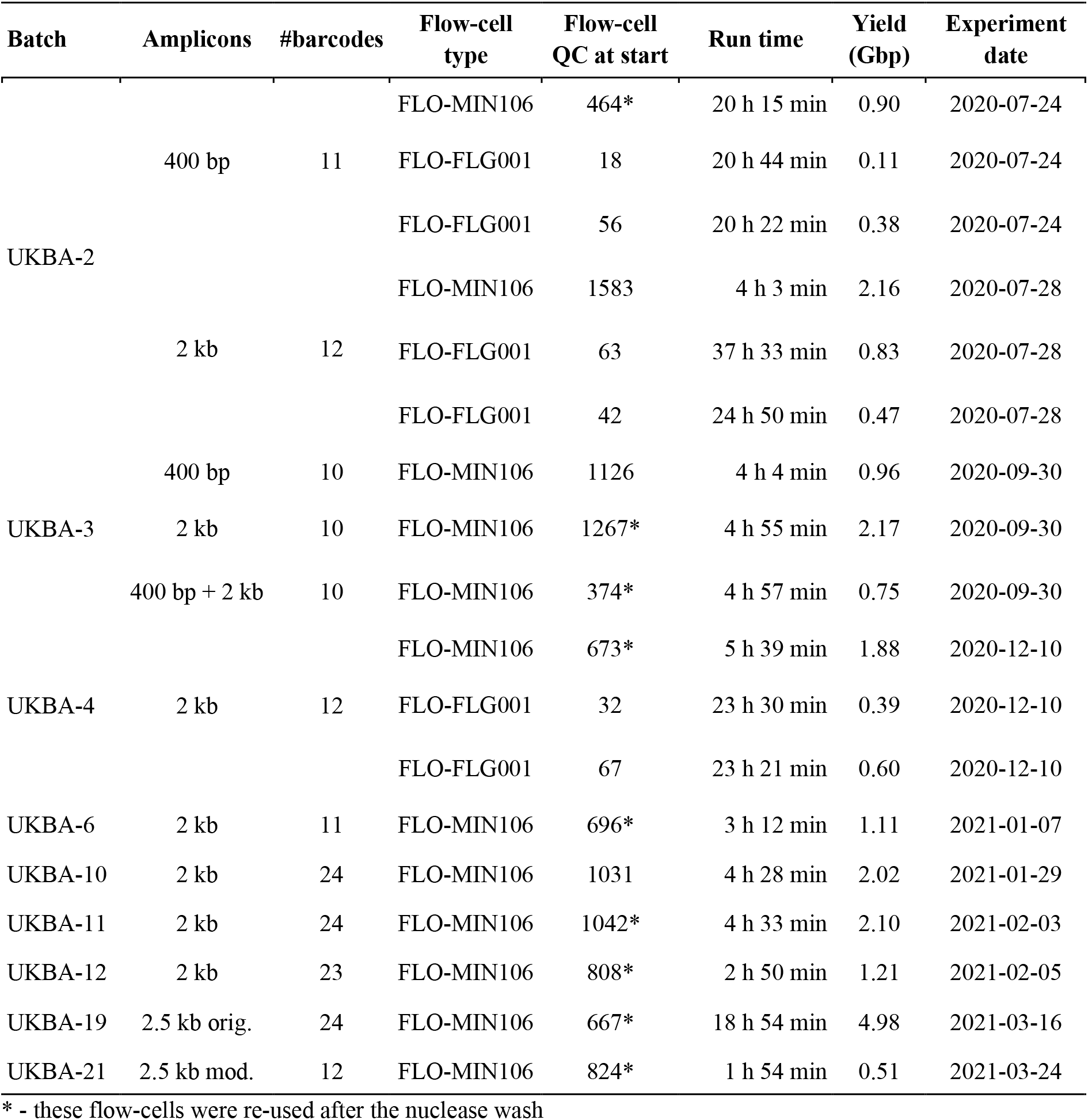
Overview of the MinION sequencing runs.

**Fig 1.**
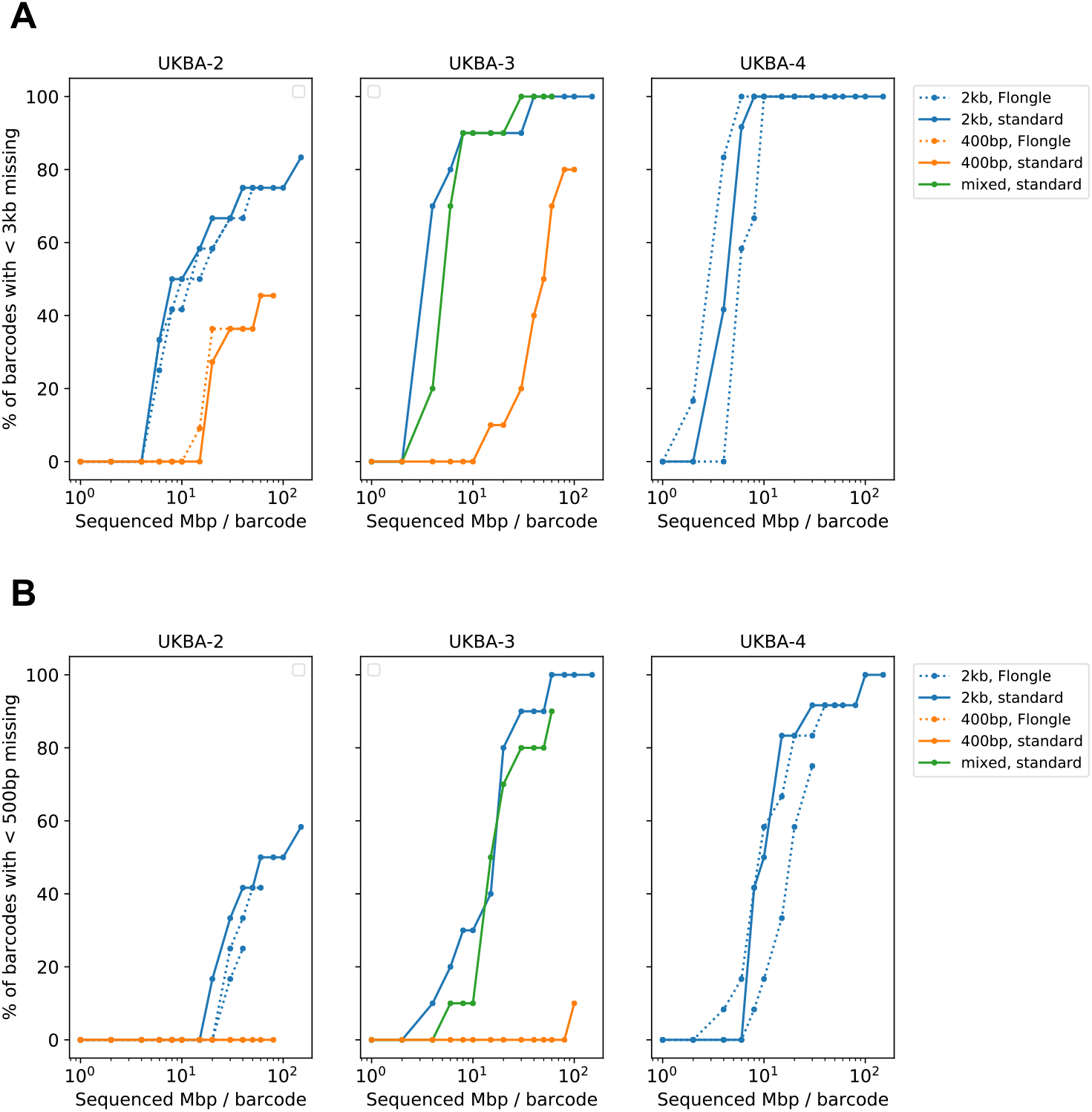
The fraction of successfully sequenced multiplexed samples over time. A sample is considered as successfully sequenced if the resulting sequence produced by the Artic pipeline has fewer than 500-bp (a) or 3-kb (b) marked as missing bases. Each run is represented by several time points, each point showing the percentage of successfully sequenced barcodes (y-axis) upon reaching a specified amount of sequenced data per barcode (x-axis).

Note that batch UKBA-2 included samples with low product concentrations after PCR amplification. As a result, three samples (barcodes 02, 06 and 11) could not be completed reliably even after combining data from all six sequencing runs. Batches UKBA-3 and UKBA-4 contained only samples with Cq values from RT-qPCR below 26. **S1 Figure** shows the amount of missing sequence in individual samples plotted against possible explanatory variables, namely the Cq values, amplicon concentration, and RNA sample storage time prior to amplification. Although the expected trends are in some cases observable, they are not followed universally.

Using the Artic pipeline for further analysis, sequencing reads must first pass a series of filters to ensure no barcode bleeding and to remove possible contamination. The number of reads passing these filters and used for the identification of variants in the final step of the pipeline varied between runs. In our experiments their fraction comprise between 14 and 55% (**Fig 2A**). Majority of failed reads are due to the low quality or incompleteness (groups A-C) and comprise between 41-78% of all reads. While there are no clear differences between short and long amplicon protocols, with 2-kb amplicons these low-quality reads are apparently more prevalent on the Flongle runs compared to the standard flow cells.

**Fig 2.**
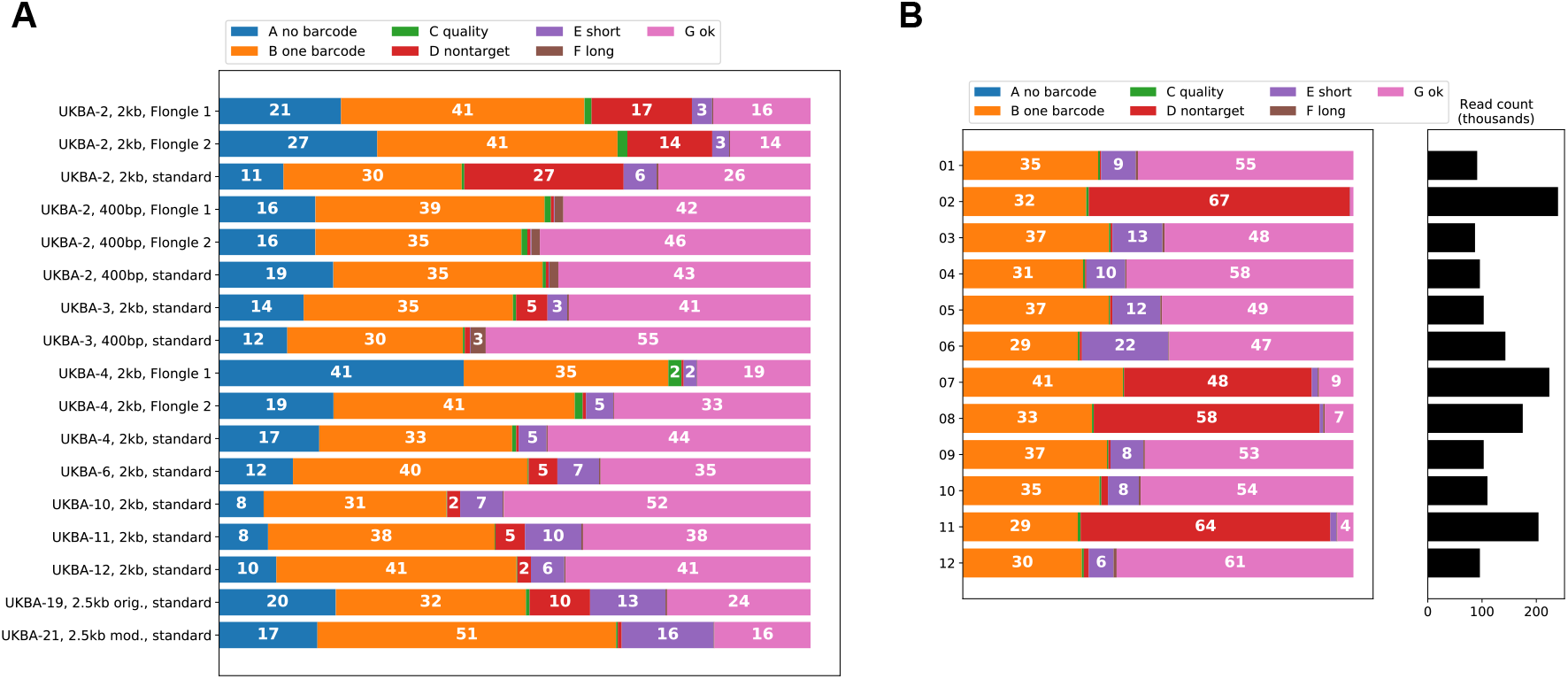
Reasons for discarding reads in the Artic pipeline. The sequencing reads must pass through a series of filters to ensure correct sample assignment and the read quality. A: reads without barcode identification. B: reads with only one barcode (Artic pipeline requires barcodes on both ends to ensure that the whole read was sequenced and to decrease the probability of barcode bleeding). C: low-quality reads (base caller quality less than 7). D: reads do not align to the SARS-CoV-2 reference. E: reads are too short (likely due to fragmentation) or F: too long (i.e. chimeric reads); lengths between 1500 and 3000 for 2-kb amplicons, between 350 and 619 for 400-bp amplicons. The reads passing all filters are included in group G. Panel (A): Summary per run. Panel (B): Detailed per-barcode analysis for UKBA-2 samples, 2-kb amplicons, standard flow cell.

Interestingly, in some runs, up to 6% of reads that pass the base quality filters do not map to the target reference genome. In particular, four samples in batch UKBA-2 of 2-kb amplicon run (barcodes 02, 07, 08 and 11) have a very high fraction of non-target reads (**Fig 2B**). The majority (82-96%) of these reads map to the human genome, and a smaller fraction (0.3-9%) map to bacterial genomes, including the species colonizing human oral cavity and respiratory tract (*e*.*g*., *Actinomyces graevenitzii, Haemophilus parainfluenzae, Leptotrichia spp*., *Prevotella spp*., *Pseudomonas aeruginosa, Rothia mucilaginosa, Streptococcus pneumoniae, S. mitis, S. parasanguinis, S. salivarius, Tannerella forsythia, Veillonella parvula*). All four samples showed a lower viral load (*i*.*e*., Cq value > 30) in RT-qPCR assays, and the amplification in the PCR-tiling protocol resulted in lower product yield. Human and bacterial reads represent artefacts apparently resulting from a non-specific amplification of contaminating nucleic acids present in clinical samples.

We have also observed that some amplicons originate from sub-genomic RNAs that co-purify with the SARS-CoV-2 genomic RNA. It has been demonstrated that the amount of sub-genomic RNAs correlates with the disease severity. As these molecules are strongly repressed in asymptomatic patients (38), their proportion in the sequencing data can serve as a molecular marker. The most abundant reads are derived from the N mRNA (39). The sub-genomic RNAs are generated in the process of the virus replication/transcription (5) and start with a leader sequence originating from the untranslated 5’ end of the viral genome, followed by a downstream sequence containing a particular open reading frame. The leftmost primer in both 400-bp and 2-kb primer sets investigated in this study is contained within the leader sequence. This facilitates amplification of sub-genomic RNAs with appropriate right primers (**Fig 3**).

**Fig 3.**
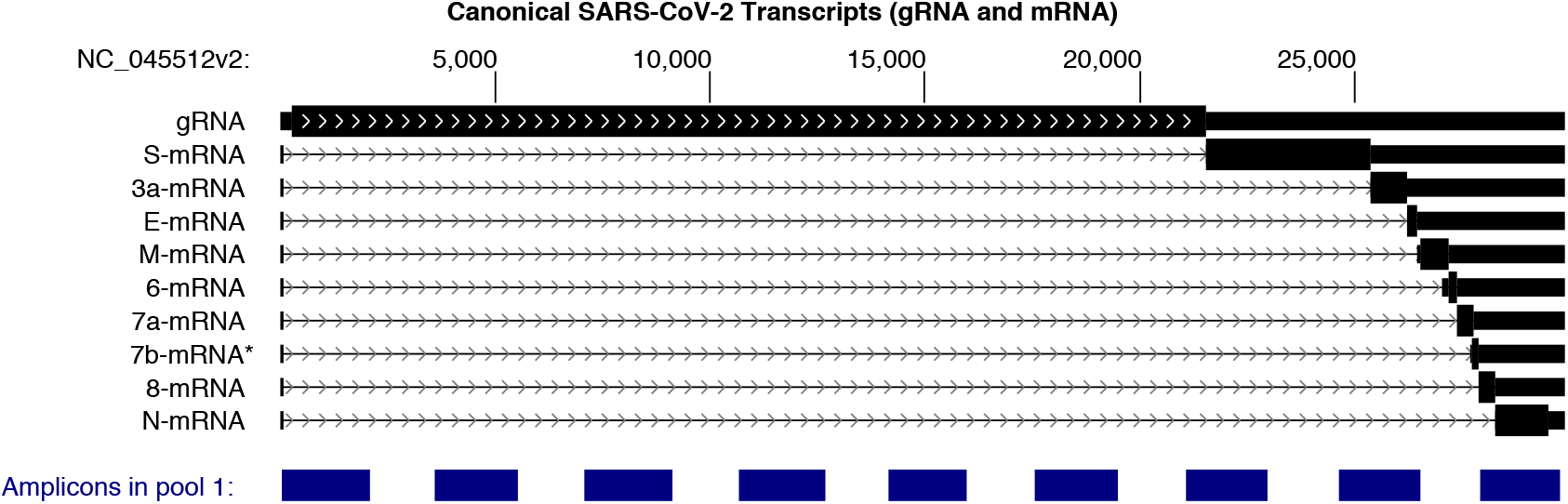
Sub-genomic RNAs. (black) and amplicons of primer pool 1 from the 2-kb primer set (blue), visualized by the UCSC genome browser (40).

**Table 2** lists the fraction of selected sub-genomic RNAs among reads that could be aligned to the SARS-CoV-2 genome. These fractions are relatively low, with the remaining sub-genomic RNAs being even more rare. However, the fractions vary among the samples. In UKBA-2 run with 2-kb amplicons, the highest fraction of 14.3% was observed for the gene N mRNA in barcode 07 and the fraction of 7.5% was observed for the ORF3a mRNA in barcode 11. Some of these sub-genomic amplicons are discarded from the analysis as too short, while others lead to lower coverage in areas not covered by the sub-genomic RNA (**Fig 4**).

**Table 2.** Sub-genomic RNA fractions. (in %) among different MinION runs with the standard flow cells (A) and in different barcodes of batch UKBA-2, 2-kb amplicons (B). Only reads that align to the SARS-CoV-2 genome and can be demultiplexed were considered. Only genes with the highest numbers of sub-genomic RNA reads are shown.

| batch | amplicon size | ORF3a | Gene E | Gene M | Gene N | Gene S | genome |
| --- | --- | --- | --- | --- | --- | --- | --- |
| **UKBA-2** | 2 kb | 2.4 | 2.5 | 1.4 | 3.5 | 0.8 | 89.3 |
|  | 400 bp | 0.0 | 0.0 | 0.1 | 0.5 | 0.0 | 99.3 |
| **UKBA-3** | 2 kb | 1.0 | 1.2 | 1.1 | 1.5 | 0.6 | 94.6 |
|  | 400 bp | 0.0 | 0.0 | 0.1 | 0.2 | 0.0 | 99.7 |
| **UKBA-4** | 2 kb | 0.9 | 0.9 | 1.0 | 1.0 | 0.8 | 95.3 |

| batch | barcode # | ORF3a | Gene E | Gene M | Gene N | Gene S | genome |
| --- | --- | --- | --- | --- | --- | --- | --- |
| **UKBA-2 (2 kb)** | 1 | 0.9 | 1.8 | 0.9 | 1.5 | 0.5 | 94.4 |
|  | 2 | 0.3 | 0.3 | 1.0 | 0.4 | 0.1 | 97.8 |
|  | 3 | 2.4 | 2.7 | 1.8 | 3.7 | 1.5 | 87.9 |
|  | 4 | 2.8 | 2.6 | 1.2 | 2.2 | 1.0 | 90.3 |
|  | 5 | 2.0 | 2.5 | 2.0 | 2.1 | 0.8 | 90.6 |
|  | 6 | 4.5 | 5.0 | 2.0 | 5.4 | 0.0 | 83.1 |
|  | 7 | 4.0 | 0.5 | 0.9 | 14.3 | 3.0 | 77.3 |
|  | 8 | 3.2 | 3.4 | 0.1 | 0.0 | 0.0 | 93.2 |
|  | 9 | 1.3 | 2.2 | 1.7 | 2.5 | 1.3 | 91.0 |
|  | 10 | 0.9 | 1.8 | 1.2 | 2.2 | 0.3 | 93.7 |
|  | 11 | 7.5 | 0.0 | 0.9 | 3.5 | 0.0 | 87.9 |
|  | 12 | 2.1 | 1.5 | 0.5 | 4.0 | 1.3 | 90.5 |

**Fig 4.**
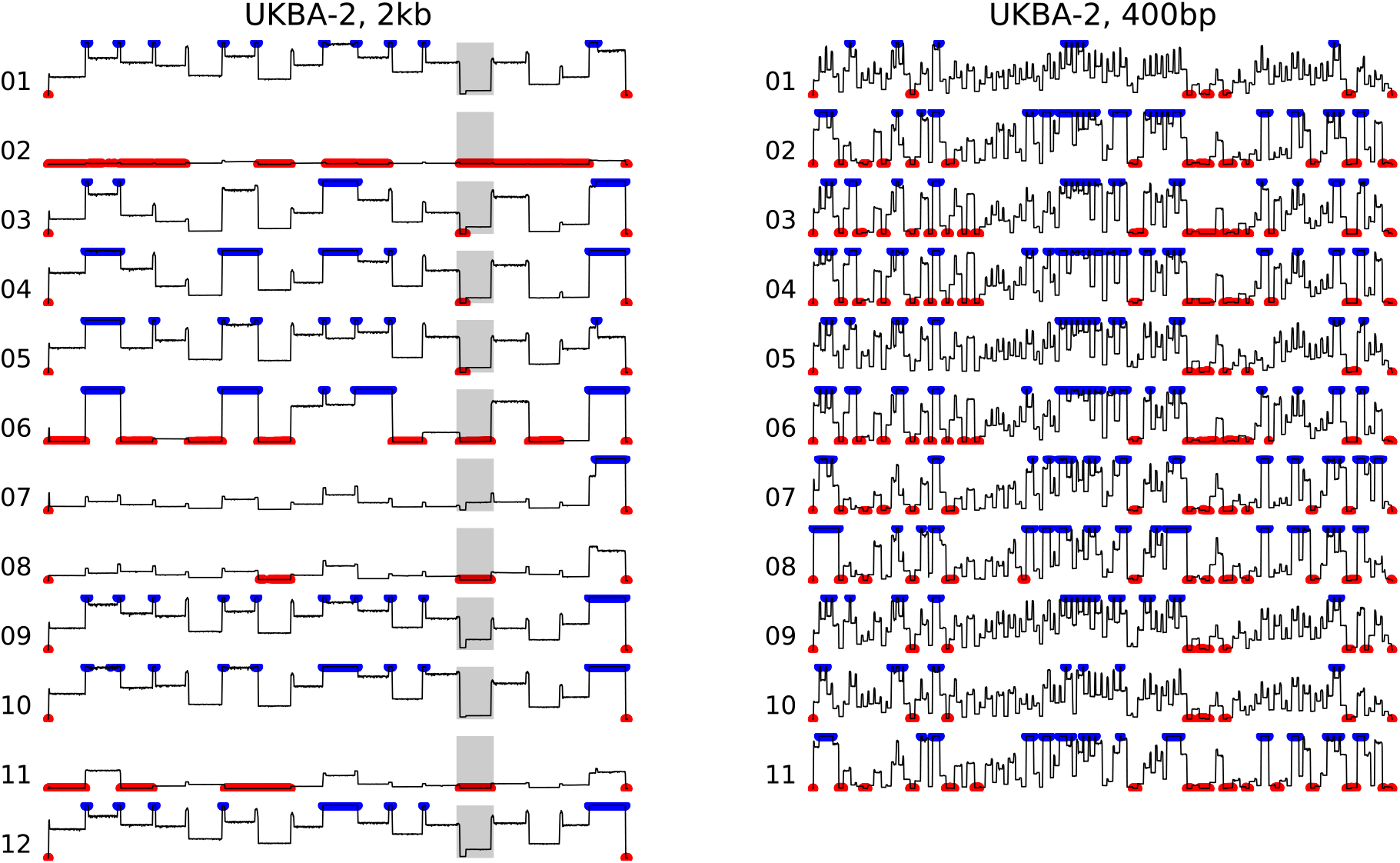
Coverage along the genome in two MinION runs for batch UKBA-2. In both runs, an initial portion of the run containing on average 40-Mbp of sequencing data per barcode was used. Coverage values higher than 1000 were clipped at this value and are shown in blue. Coverage below 20 (default Artic cutoff) is shown in red. Medians of 10-bp windows are shown for smoothing. The very starts and ends of the genome are not covered by amplicons, and are thus displayed in red. Shaded area in the left column corresponds to amplicon 13. Some barcodes have a visible dip in the coverage at the left end of this amplicon; this difference in coverage is caused by reads originating from sub-genomic RNAs corresponding to the gene S. Similar plots for additional runs are shown in **S2 Fig**.

From these pilot experiments, we conclude that even though 400-bp amplicons have a lower percentage of discarded reads (**Fig 2**), they produce fewer finished sequences at a comparable overall amount of sequence data (**Fig 1**). The reason is a very uneven coverage of individual amplicons (**Fig 4**). This is observed in both sets of primers, but for the 400-bp amplicons we see a much lower coverage in the worst covered regions (**Fig 5**). Additional sequencing runs (UKBA-6, UKBA-10, UKBA-11, and UKBA-12) were performed with long 2-kb amplicons on standard MinION flow cells with similar results (**Fig 2A; S2 Fig**).

**Fig 5.**
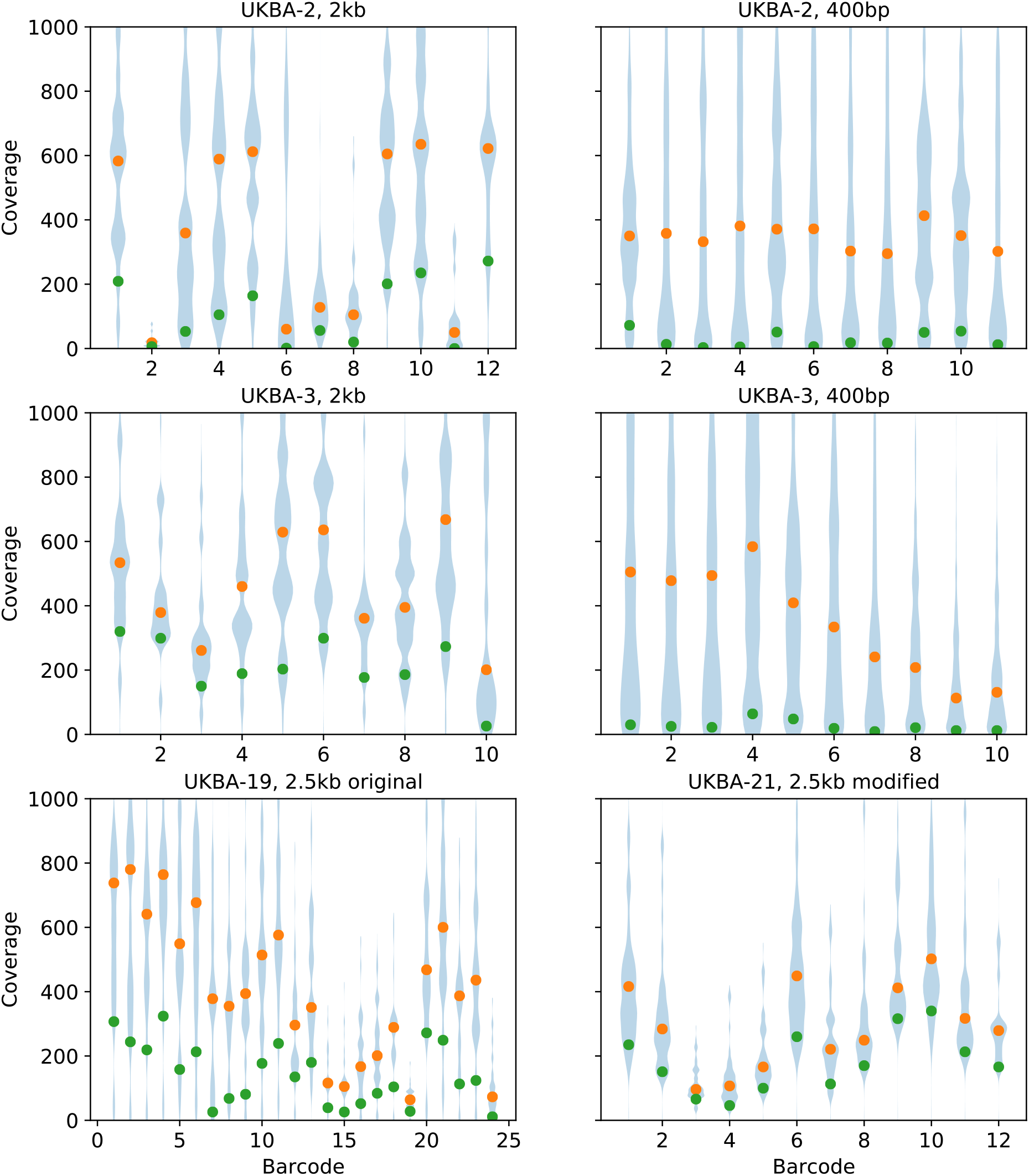
Coverage distribution in different sequencing runs. For each barcode, coverage by reads passing the Artic filter was computed along the genome and the distribution of the coverage values was summarized as a violin plot (blue), cropped at coverage 1000. Orange dots represent median coverage and green dots 10% percentile (corresponding to a approx. 3-kb portion of the genome with the lowest coverage). In all runs, an initial portion containing on average 40-Mbp of sequencing data per barcode was used.

To investigate if a different primer scheme for generating long amplicons can solve the problem with uneven coverage (in particular, see amplicon 13 in **Fig 4** which partially covers the S gene region important for identification of the SARS-CoV-2 Variants of Concerns), we also tested the 2.5-kb primer panel (27). Except for the leftmost primer, the primer positions in this panel differ from those of the 2-kb scheme. We have performed two sequencing runs with 2.5-kb primer set (UKBA-19, UKBA-21). In the first experiment, we have noticed an almost complete drop of coverage in the last amplicon derived from the 3’ end of the genome; for the second experiment, we have replaced the primers for the right-most amplicon with the right-most primers from the 2-kb panel, which mitigated the issue. Comparing the coverage of individual amplicons between the 2-kb and 2.5-kb schemes (**Fig 6**), the coverage in the 2.5-kb scheme indeed appears to be more even. **Fig 5** illustrates that our modification of the 2.5-kb scheme leads to a particularly small difference between the median coverage and coverage of the lowest 10% of the genome, which may result in fewer regions with insufficient coverage. However, we have also noticed a higher percentage of failed reads, with only 24% (UKBA-19) and 16% (UKBA-21) reads passing all filters and being usable for variant identification. Further analysis revealed a notable increase in single-barcode reads (group B) and shorter than expected reads (group E), pointing to difficulties in amplifying and sequencing longer fragments. More experiments are required to determine whether the 2.5-kb scheme results in more fully-assembled genomes over the 2-kb scheme.

**Fig 6.**
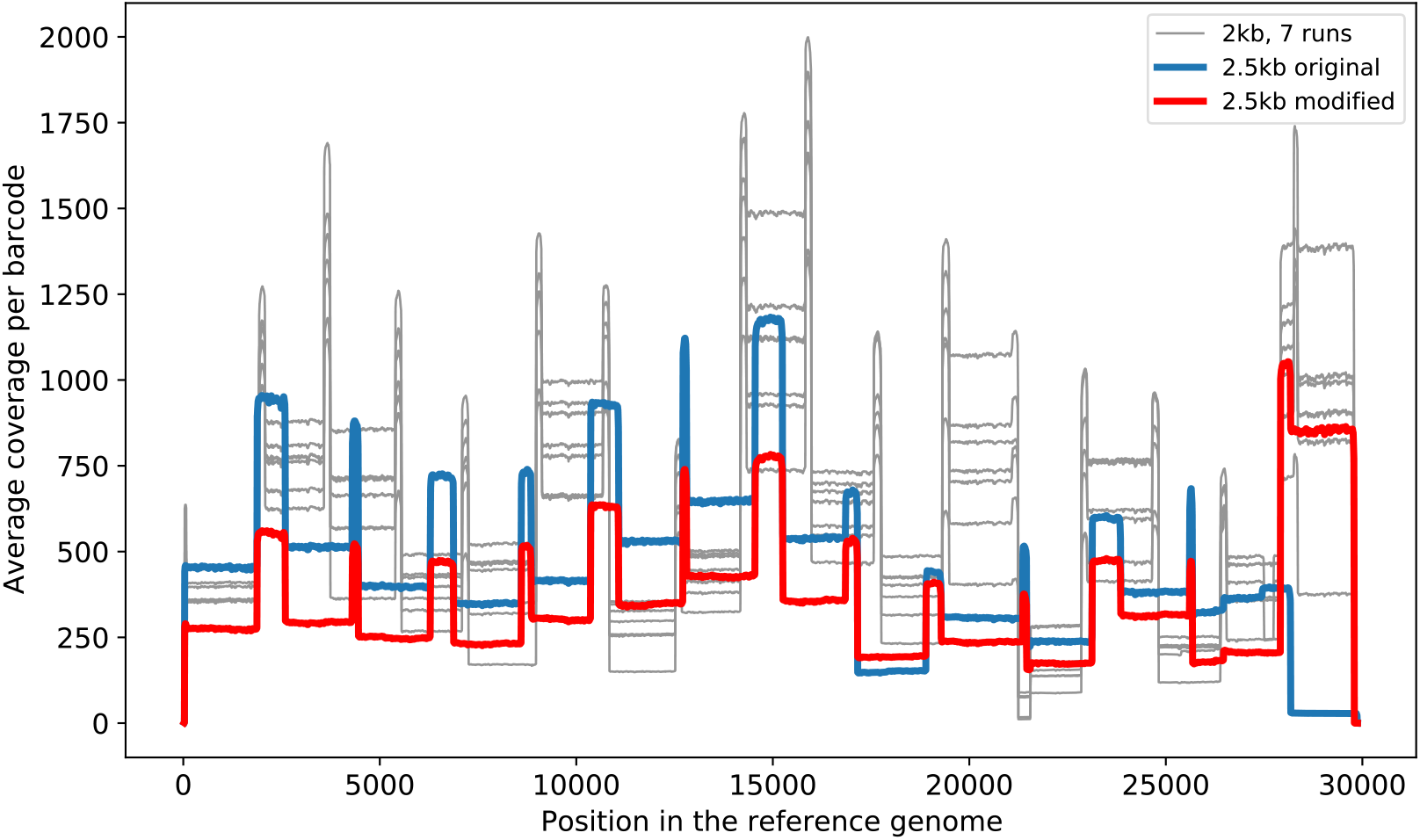
Average coverage along the genome for seven runs with 2-kb amplicons (batches UKBA-2,3,4,6,10,11,12) and two runs with 2.5-kb amplicons (UKBA-19 with the original primer set and UKBA-21 with the last primer pair replaced by its counterpart from the 2-kb scheme). Each line depicts the average coverage over all samples in a run at the time point when 40-Mbp per sample was sequenced on average. Medians of 50-bp windows are shown for smoothing. Note a drop-out in the amplicon 13 (2-kb scheme) which covers a 3’ end of orf1b and about a third of the S gene including the region associated with mutations in Variants of Concern such as B.1.1.7.

## Conclusions

In this paper, we have compared three versions of PCR-tiling protocol for sequencing SARS-CoV-2 genomes from clinical samples on MinION platform. Our results have shown that even though the protocol based on short 400-bp amplicons generally produces more usable data, the coverage of individual amplicons varies widely which may result in difficulties in recovering individual mutations in under-represented amplicons. In comparison, longer amplicons tend to produce close-to-finished genomes more quickly, generally requiring smaller amounts of raw data produced per barcode sequenced. However, protocols based on long amplicons produce a higher percentage of reads that are unsuitable for further analysis with the Artic pipeline, likely due to a combination of fragmentation of synthesized molecules and prematurely aborted molecules during sequencing. The longer amplicon protocols are also less suitable for applications, where original RNA molecules in clinical samples may already be fragmented. Generally, the Flongle flow cells performed worse in sequencing multiplexed libraries containing barcoded samples than regular MinION flow cells, which have an added advantage of ability to adjust the length of the run based on the library and individual sample quality.

Interestingly, PCR-tiling protocols were able to also pick up sub-genomic RNA transcripts, and the proportion of these transcripts varied between samples. Since increased levels of sub-genomic transcripts are correlated with severe cases of COVID-19, these protocols could be optimized to detect the levels of sub-genomic transcripts more accurately and used as a biomarker for disease severity.

It is evident that effective epidemiologic surveillance of the pandemic is strongly dependent on systematic sequencing of SARS-CoV-2 isolates. The MinION platform from Oxford Nanopore Technologies is one of the most powerful and versatile means for acquisition of viral sequences. Yet, as demonstrated in this study, the pros and cons of a particular protocol must be taken into account to ensure that the sequencing results will be of a highest quality, which is an essential prerequisite for their utility in fighting the pandemic.

## Materials and Methods

### Collection of samples and RNA preparation

Oropharyngeal swabs of patients with suspected COVID-19, collected between March 30, 2020 and March 19, 2021, were preserved in 2-3 ml of viral transport medium and delivered to the laboratory of the Biomedical Research Centre of the Slovak Academy of Sciences in Bratislava, Slovakia in frame of the routine RT-qPCR diagnostics for SARS-CoV-2. Initially (UKBA-2 samples), 100 µl of the swab medium was used for the RNA extraction using the Zymo Research Quick-RNA™ Viral 96 Kit (Zymo Research, Irvin, California, USA). Resulting RNA was eluted to 20 µl of nuclease free water. For all other specimens, the Biomek i5 Automated Workstation (Beckman Coulter, Indianapolis, Indiana, USA) was employed using the RNAdvanced Viral kit (Beckman Coulter, Indianapolis, Indiana, USA). In this case, RNA was extracted from 200 µl of swab medium and eluted to 40 µl of nuclease free water.

### Real-time quantitative PCR (RT-qPCR)

In frame of the routine RT-qPCR diagnostics, presence of SARS-CoV-2 RNA was detected by vDETECT COVID-19 RT-qPCR kit, rTEST COVID-19 RT-qPCR kit or rTEST COVID-19 RT-qPCR ALLPLEX kit (MultiplexDX, Bratislava Slovakia) targeting RNA-dependent RNA polymerase (RdRp) and Envelope (E) genes. The RT-qPCR assays were carried on QuantStudio(tm) 5 Real-Time PCR System (Applied Biosystem, Foster City, California, USA).

### Library preparation and DNA sequencing

The sequencing libraries were constructed using a ligation kit (SQK-LSK109) essentially as described in a PCR-tiling of COVID-19 virus protocol (PTC_9096_v109_revF_06Feb2020; Oxford Nanopore Technologies) with minor modifications. Briefly, RNA samples extracted from swabs positive for the presence of SARS-CoV-2 in RT-qPCR assay (quantification cycle (Cq) values 13.46 - 32.03; **S1 Table**) were converted into cDNA using a SuperScript IV reverse transcriptase (Thermo Fisher Scientific) or LunaScript® RT SuperMix Kit (New England Biolabs). For each sample, the overlapping amplicons were generated using a Q5® Hot Start High-Fidelity DNA polymerase (New England Biolabs) and the primer pools spanning the SARS-CoV-2 genome sequence (*i*.*e*., 400-bp Artic nCoV-2019 V3 panel (https://github.com/artic-network/artic-ncov2019) purchased from Integrated DNA Technologies (IDT, cat.no. 10006788) and the 2-kb (35) and 2.5-kb schemes (27), custom synthesized by Microsynth). The same cycling program was used for all amplicon types (*i*.*e*., 30 sec initial denaturation at 98°C, followed by 25 to 35 cycles of 15 sec at 98°C (denaturation) and 5 min at 65°C (combined annealing and polymerization), and cooling to 4°C). The amplifications were performed in two separate reactions and the overlapping amplicons were pooled, purified using an equal volume of AMPure XP magnetic beads (Beckman Coulter) and quantified using a Qubit 3.0 spectrophotometer and dsDNA Broad Range Assay Kit (Thermo Fisher Scientific). About 50-75 ng (400-bp amplicons) and 250-300 ng (2 and 2.5-kb amplicons) of each SARS-CoV-2 isolate were treated with NEBNext Ultra II End repair / dA-tailing Module (New England Biolabs). The samples were then barcoded using EXP-NBD104 (barcodes 1-12) or EXP-NBD114 (barcodes 13-24) kits (Oxford Nanopore Technologies) and NEBNext Ultra II Ligation Master Mix (New England Biolabs). Barcoded samples were pooled and purified using 0.6 volume of AMPure XP magnetic beads. The AMII sequencing adapter (Oxford Nanopore Technologies) was ligated to about 75 ng (400-bp amplicons) or 300 ng (2 and 2.5-kb amplicons) of barcoded pools using Quick T4 DNA ligase (New England Biolabs) and the sequencing libraries were purified using 0.6 volume of AMPure XP magnetic beads. About 20 ng (400-bp amplicons) and 90 ng (2 and 2.5-kb amplicons) of the libraries were loaded on an R9.4.1 flow cell (FLO-MIN106). The sequencing was performed using a MinION Mk-1b device (Oxford Nanopore Technologies). For sequencing on the Flongle flow cells (FLO-FLG001), the library preparation was the same, except that one third to one half of the library was loaded compared to the amount used for the standard flow cell.

### Data processing

Nanopore sequencing data were base called and demultiplexed using Guppy v.3.4.4. Variant analysis was performed using Artic analysis pipeline v.1.1.3. (https://artic.network/ncov-2019/ncov2019-bioinformatics-sop.html) using recommended settings. Minimum and maximum read lengths in the Artic guppyplex filter were set to 350 and 619 for the 400-bp amplicons and 1500 and 3000 for both the 2 and 2.5-kb amplicons, respectively. For batch UKBA-2, the final sequences were produced by first combining sequencing reads from both standard and Flongle runs with the same primer set and running the Artic pipeline. Subsequently the results for the two primer sets were combined so that regions sufficiently covered by at least one amplicon set were considered as finished. The same process was used in batch UKBA-3, but there was only data from standard flow cells available. Subsequent batches were based on 2 or 2.5-kb amplicons sequenced on a standard flow cell.

To compare different primer sets and sequencing devices, reads were also demultiplexed at the less strict default Guppy settings and aligned to various reference genomes by minimap2 v. 2.13-r852-dirty (41). Reference genomes include the SARS-CoV-2 genome MN908947.3 (1), the human genome version hg19 downloaded from the UCSC genome browser (40), and the database for bacterial species typing included in the Japsa software (42). To detect sub-genomic RNAs, reads were aligned to transcripts downloaded from the UCSC genome browser by minimap2, and classified as sub-genomic, if the alignment to a sub-genomic RNA has at least 5 matches more than the best alignment to the reference genome. Read coverage was computed using genomecov tool from BEDTools (43) with options -bga -split.

To compare the results for various sequencing data volumes, reads were ordered by the sequencing finish time and the initial portion with the desired total length was selected and used for the analysis in the Artic pipeline. To compare batches with a different number of samples, the cutoffs were expressed as the average amount per barcode.

### Data availability

The SARS-CoV-2 genome sequences determined in this study were deposited in the GISAID database (http://www.gisaid.org). The accession IDs are shown in **S1 Table**. The sequencing reads mapping to the reference genome were deposited to ENA under study PRJEB44303.

## Data Availability

The SARS-CoV-2 genome sequences determined in this study were deposited in the GISAID database (http://www.gisaid.org). The accession IDs are shown in S1 Table. The sequencing reads mapping to the reference genome were deposited to ENA under study PRJEB44303.

## Acknowledgements

The authors wish to thank Lubomir Tomaska (Comenius University in Bratislava) for critical reading of the manuscript and discussions. The project was supported by grants from the Slovak Research and Development Agency (APVV-18-0239 to JN, PP-COVID-20-0017 to BK), the Scientific Grant Agency (VEGA 1/0463/20 to BB, VEGA 1/0458/18 to TV, VEGA 1/0027/19 to JN, VEGA 1/0136/20 to MN), and the European Union’s Horizon 2020 research and innovation program (EVA-GLOBAL project #871029 to BK and PANGAIA project #872539). The research was also supported in part by the Operation Program of Integrated Infrastructure (OPII) projects ITMS2014: 313011ATL7 and ITMS2014+: 313021×329 (Advancing University Capacity and Competence in Research, Development and Innovation), co-financed by the European Regional Development Fund.

## Ethics statement

All clinical specimens used within this study were previously collected for the purpose of primary diagnosis of SARS-CoV-2 and were transferred to the Biomedical Research Center of the Slovak Academy of Sciences, Bratislava, Slovakia while made unidentifiable for the researchers performing this study. The study has been approved by the Ethics committee of Biomedical Research Center of the Slovak Academy of Sciences, Bratislava, Slovakia (Ethics committee statement No. EK/BmV-02/2020).

## Author contributions

Conceived and designed the experiments: BB, JN, TV. Performed the experiments: KB, VC, DF, VH, JN. Analyzed the data: BB, AG, TV. Wrote the first draft of manuscript: BB, JN, TV. Edited and approved the manuscript: BB, KB, VC, DF, AG, VH, BK, MN, JN, TV. Obtained funding: BB, BK, MN, JN, TV.

## Competing interests

The authors declare that they have no conflict of interest.

## Notes

### Competing Interest Statement

The authors have declared no competing interest.

