## Supporting Information for "Nanopore Sequencing of SARS-CoV-2: Comparison of Short and Long PCR-tiling Amplicon Protocols"

#### Supporting Information Captions

**S1 Table. Overview of the SARS-CoV-2 samples sequenced in this study.**

**S1 Fig. Dependence of the amount of missing sequence after Artic analysis on various sample properties**. (A) Cq value of the diagnostic RT-qPCR test, (B) DNA concentration after amplification (C) length of storage of the sample before PCR amplification. Each dot corresponds to one sample, each sub-plot has a different level of sequencing per barcode.

**S2 Fig. Coverage along the genome in several MinION runs**. In all runs, an initial portion of the run containing on average 40-Mbp of sequencing data per barcode was used. Coverage values higher than 1000 were clipped at this value and are shown in blue. Coverage below 20 (default Artic cutoff) is shown in red. Medians of 10-bp windows are shown for smoothing.

| **S1 Table.** Overview of the SARS-CoV-2 samples sequenced in this study. | | | | | | | |
|---|---|---|---|---|---|---|---|
| **Batch** | **Sample ID** | **Sample type** | **Location [Slovakia]** | **RT-qPCR [Cq value]** | **Barcode** | **GISAID ID** | **Pangolin lineage [https://pangolin.cog-uk.io]** |
| UKBA-2 | UKBA-201/2020 | swab | Martinček | 21.36 | 01 | EPI_ISL_577734 | B.1.1.257 |
| | UKBA-202/2020 | swab | Bratislava | 31.06 | 02 | not submitted | n/a |
| | UKBA-203/2020 | swab | Žehra | 19.34 | 03 | EPI_ISL_577735 | B.1.1 |
| | UKBA-204/2020 | swab | Bratislava | 22.15 | 04 | EPI_ISL_577736 | B.1.1 |
| | UKBA-205/2020 | swab | Žehra | 18.40 | 05 | EPI_ISL_577737 | B.1.1 |
| | UKBA-206/2020 | swab | Žehra | 20.57 | 06 | not submitted | n/a |
| | UKBA-207/2020 | swab | Pezinok | 30.80 | 07 | EPI_ISL_577738 | B.1.1 |
| | UKBA-208/2020 | swab | Pezinok | 32.03 | 08 | EPI_ISL_577739 | B.1.1 |
| | UKBA-209/2020 | swab | Bratislava | 22.09 | 09 | EPI_ISL_577740 | B.1.1.70 |
| | UKBA-210/2020 | swab | Bratislava | 18.46 | 10 | EPI_ISL_577741 | B.1.131 |
| | UKBA-211/2020 | swab | Bratislava | 31.49 | 11 | not submitted | n/a |
| | UKBA-212/2020 | swab | Bratislava | 24.36 | 12 | EPI_ISL_577742 | B.1.1 |
| UKBA-3 | UKBA-313/2020 | swab | Tvrdošovce | 25.58 | 13 | EPI_ISL_583481 | B.1.1.70 |
| | UKBA-314/2020 | swab | Jatov | 25.37 | 14 | EPI_ISL_583482 | B.1.160 |
| | UKBA-315/2020 | swab | Kameničná | 23.12 | 15 | EPI_ISL_583483 | B.1.160 |
| | UKBA-316/2020 | swab | Bratislava | 18.05 | 16 | EPI_ISL_583484 | B.1.160 |
| | UKBA-317/2020 | swab | Šurany | 17.87 | 17 | EPI_ISL_583485 | B.1.160 |
| | UKBA-318/2020 | swab | Šurany | 24.28 | 18 | EPI_ISL_583486 | B.1.1.170 |
| | UKBA-319/2020 | swab | Bratislava | 26.67 | 19 | EPI_ISL_583487 | B.1.1.70 |
| | UKBA-320/2020 | swab | Bratislava | 20.50 | 20 | EPI_ISL_717975 | B.1.1.273 |
| | UKBA-321/2020 | swab | Bratislava | 18.11 | 21 | EPI_ISL_583488 | B.1.5 |
| | UKBA-322/2020 | swab | Levice | 16.64 | 22 | EPI_ISL_583489 | B.1.1.70 |
| UKBA-4 | UKBA-401/2020 | swab | Bratislava | 21.03 | 01 | EPI_ISL_779403 | B.1.1.163 |
| | UKBA-402/2020 | swab | Bratislava | 17.90 | 02 | EPI_ISL_718250 | B.1.160 |
| | UKBA-403/2020 | swab | Bratislava | 20.22 | 03 | EPI_ISL_718251 | B.1.258 |
| | UKBA-404/2020 | swab | Bratislava | 16.50 | 04 | EPI_ISL_718252 | B.1.258 |
| | UKBA-405/2020 | swab | Bratislava | 17.24 | 05 | EPI_ISL_718253 | B.1.258 |
| | UKBA-406/2020 | swab | Bratislava | 14.54 | 06 | EPI_ISL_718254 | B.1.160 |
| | UKBA-407/2020 | swab | Bratislava | 17.41 | 07 | EPI_ISL_718255 | B.1.160 |
| | UKBA-408/2020 | swab | Bratislava | 17.72 | 08 | EPI_ISL_718256 | B.1.160 |
| | UKBA-409/2020 | swab | Nitra | 22.72 | 09 | EPI_ISL_718257 | B.1.1.170 |
| | UKBA-410/2020 | swab | Nitra | 17.62 | 10 | EPI_ISL_718258 | B.1 |
| | UKBA-411/2020 | swab | Bratislava | 18.89 | 11 | EPI_ISL_718259 | B.1.1 |
| | UKBA-412/2020 | swab | Nové Zámky | 19.16 | 12 | EPI_ISL_718260 | B.1.1.170 |
| UKBA-6 | UKBA-601/2020 | swab | Nové Zámky | 17.54 | 01 | EPI_ISL_788979 | B.1.160 |
| | UKBA-602/2020 | swab | Nové Zámky | 17.95 | 02 | EPI_ISL_788980 | B.1 |
| | UKBA-603/2020 | swab | Nové Zámky | 20.21 | 03 | EPI_ISL_788981 | B.1 |
| | UKBA-604/2020 | swab | Bratislava | 20.05 | 04 | EPI_ISL_788982 | B.1.258 |
| | UKBA-605/2020 | swab | Doľany | 24.00 | 05 | EPI_ISL_791990 | B.1.1.163 |
| | UKBA-606/2020 | swab | Bratislava | 24.17 | 06 | EPI_ISL_788983 | B.1.160 |
| | UKBA-607/2020 | swab | Bratislava | 20.65 | 07 | EPI_ISL_788984 | B.1.160 |
| | UKBA-608/2020 | swab | Miloslavov | 23.30 | 08 | EPI_ISL_788985 | B.1.258 |
| | UKBA-609/2020 | swab | Bratislava | 20.37 | 09 | EPI_ISL_788986 | B.1.160 |
| | UKBA-610/2020 | swab | Bratislava | 21.59 | 10 | EPI_ISL_788987 | B.1.160 |
| | UKBA-611/2020 | swab | Bratislava | 25.67 | 11 | EPI_ISL_788988 | B.1.160 |
| UKBA-10 | UKBA-1001/2020 | swab | Trenčín | 15.00 | 01 | EPI_ISL_903980 | B.1.258 |
| | UKBA-1002/2020 | swab | Trenčín | 20.20 | 02 | EPI_ISL_903981 | B.1.1.170 |
| | UKBA-1003/2020 | swab | Trenčín | 22.70 | 03 | EPI_ISL_903982 | B.1.258 |
| | UKBA-1004/2020 | swab | Trenčín | 14.90 | 04 | EPI_ISL_903983 | B.1.258 |
| | UKBA-1005/2020 | swab | Trenčín | 20.20 | 05 | EPI_ISL_903984 | B.1.258 |
| | UKBA-1006/2020 | swab | Trenčín | 13.90 | 06 | EPI_ISL_903985 | B.1.258 |
| | UKBA-1007/2020 | swab | Trenčín | 13.70 | 07 | EPI_ISL_903986 | B.1.1.7 |
| | UKBA-1008/2020 | swab | Trenčín | 16.10 | 08 | EPI_ISL_903987 | B.1.258 |
| | UKBA-1009/2020 | swab | Trenčín | 17.70 | 09 | EPI_ISL_903988 | B.1.160 |
| | UKBA-1010/2020 | swab | Trenčín | 17.30 | 10 | EPI_ISL_903989 | B.1.1.7 |
| | UKBA-1011/2020 | swab | Trenčín | 21.40 | 11 | EPI_ISL_903990 | B.1.1.7 |
| | UKBA-1012/2020 | swab | Trenčín | 21.50 | 12 | EPI_ISL_903991 | B.1.258 |
| | UKBA-1013/2020 | swab | Trenčín | 14.20 | 13 | EPI_ISL_903992 | B.1.177 |
| | UKBA-1014/2020 | swab | Trenčín | 20.10 | 14 | EPI_ISL_903993 | B.1.258 |
| | UKBA-1015/2020 | swab | Trenčín | 22.70 | 15 | EPI_ISL_903994 | B.1.1.277 |
| | UKBA-1016/2020 | swab | Trenčín | 19.90 | 16 | EPI_ISL_903995 | B.1.258 |
| | UKBA-1017/2020 | swab | Trenčín | 16.70 | 17 | EPI_ISL_903996 | B.1.160 |
| | UKBA-1018/2020 | swab | Trenčín | 16.40 | 18 | EPI_ISL_903997 | B.1.258 |
| | UKBA-1019/2020 | swab | Trenčín | 15.90 | 19 | EPI_ISL_903998 | B.1.258 |
| | UKBA-1020/2020 | swab | Trenčín | 14.70 | 20 | EPI_ISL_903999 | B.1.258 |
| | UKBA-1021/2020 | swab | Trenčín | 16.90 | 21 | EPI_ISL_904000 | B.1.1.7 |
| | UKBA-1022/2020 | swab | Trenčín | 14.80 | 22 | EPI_ISL_904001 | B.1.221 |
| | UKBA-1023/2020 | swab | Trenčín | 18.70 | 23 | EPI_ISL_904002 | B.1.258 |
| | UKBA-1024/2020 | swab | Trenčín | 22.50 | 24 | EPI_ISL_904003 | B.1.258 |
| | UKBA-1101/2021 | swab | Dolný Kubín | 19.23 | 01 | EPI_ISL_959643 | B.1.1.7 |
| | UKBA-1102/2021 | swab | Dolný Kubín | 16.87 | 02 | EPI_ISL_959642 | B.1.160 |
| | UKBA-1103/2021 | swab | Dolný Kubín | 16.35 | 03 | EPI_ISL_959648 | B.1.258 |
| | UKBA-1104/2021 | swab | Dolný Kubín | 14.93 | 04 | EPI_ISL_959645 | B.1.1.7 |
| | UKBA-1105/2021 | swab | Ružomberok | 17.68 | 05 | EPI_ISL_959647 | B.1.1.170 |

| | | | | | | | |
|---|---|---|---|---|---|---|---|
| UKBA-11 | UKBA-1106/2021 | swab | Dolný Kubín | 18.64 | 06 | EPI_ISL_959646 | B.1.1.7 |
| | UKBA-1107/2021 | swab | Dolný Kubín | 21.26 | 07 | EPI_ISL_959644 | B.1.1.7 |
| | UKBA-1108/2021 | swab | Dolný Kubín | 19.74 | 08 | EPI_ISL_959649 | B.1.258 |
| | UKBA-1109/2021 | swab | Rimavská Sobota | 24.25 | 09 | EPI_ISL_959637 | B.1.1.7 |
| | UKBA-1110/2021 | swab | Rimavská Sobota | 21.76 | 10 | EPI_ISL_959638 | B.1.1.7 |
| | UKBA-1111/2021 | swab | Revúca | 16.65 | 11 | EPI_ISL_959639 | B.1.1.7 |
| | UKBA-1112/2021 | swab | Dolný Kubín | 20.69 | 12 | EPI_ISL_959640 | B.1.1.7 |
| | UKBA-1113/2021 | swab | Dolný Kubín | 22.52 | 13 | EPI_ISL_959641 | B.1.1.170 |
| | UKBA-1114/2021 | swab | Ružomberok | 23.99 | 14 | EPI_ISL_959627 | B.1.1.7 |
| | UKBA-1115/2021 | swab | Trenčianska Teplá | 23.14 | 15 | EPI_ISL_959630 | B.1.258 |
| | UKBA-1116/2021 | swab | Ružomberok | 25.20 | 16 | EPI_ISL_959628 | B.1.1.7 |
| | UKBA-1117/2021 | swab | Bratislava | 23.07 | 17 | EPI_ISL_959626 | B.1.1.7 |
| | UKBA-1118/2021 | swab | Trenčianska Teplá | 16.29 | 18 | EPI_ISL_959631 | B.1.258 |
| | UKBA-1119/2021 | swab | Košice | 17.18 | 19 | EPI_ISL_959629 | B.1.1.7 |
| | UKBA-1120/2021 | swab | Bartošova Lehôtka | 15.78 | 20 | EPI_ISL_959632 | B.1.1.7 |
| | UKBA-1121/2021 | swab | Muránska Huta | 16.60 | 21 | EPI_ISL_959633 | B.1.1.7 |
| | UKBA-1122/2021 | swab | Veľký Krtíš | 15.92 | 22 | EPI_ISL_959634 | B.1.1.7 |
| | UKBA-1123/2021 | swab | Mýto pod Ďumbierom | 15.92 | 23 | EPI_ISL_959635 | B.1.1.7 |
| | UKBA-1124/2021 | swab | Želovce | 16.75 | 24 | EPI_ISL_959636 | B.1.258 |
| UKBA-12 | UKBA-1201/2021 | swab | Dolný Kubín | 17.63 | 01 | EPI_ISL_959623 | B.1.258 |
| | UKBA-1202/2021 | swab | Dolný Kubín | 17.78 | 02 | EPI_ISL_959624 | B.1.1.7 |
| | UKBA-1203/2021 | swab | Tvrdošín | 17.64 | 03 | EPI_ISL_959625 | B.1.1.170 |
| | UKBA-1204/2021 | swab | Revúca | 15.27 | 04 | EPI_ISL_959622 | B.1.1.7 |
| | UKBA-1205/2021 | swab | Trstenná | 16.87 | 05 | EPI_ISL_1234384 | B.1.1.7 |
| | UKBA-1206/2021 | swab | Hnúšťa | 15.31 | 06 | EPI_ISL_959621 | B.1.1.7 |
| | UKBA-1207/2021 | swab | Bratislava | 16.49 | 07 | EPI_ISL_959604 | B.1.1.7 |
| | UKBA-1208/2021 | swab | Bratislava | 23.46 | 08 | EPI_ISL_959605 | B.1.1.7 |
| | UKBA-1209/2021 | swab | Bratislava | 13.46 | 09 | EPI_ISL_959606 | B.1.1.7 |
| | UKBA-1210/2021 | swab | Bratislava | 14.50 | 10 | EPI_ISL_959607 | B.1.258 |
| | UKBA-1211/2021 | swab | Bratislava | 17.59 | 11 | EPI_ISL_959608 | B.1.1.7 |
| | UKBA-1212/2021 | swab | Bratislava | 15.51 | 12 | EPI_ISL_959609 | B.1.1.7 |
| | UKBA-1213/2021 | swab | Bratislava | 21.57 | 13 | EPI_ISL_959610 | B.1.1.7 |
| | UKBA-1214/2021 | swab | Bratislava | 14.68 | 14 | EPI_ISL_959611 | B.1.1.7 |
| | UKBA-1215/2021 | swab | Bratislava | 17.21 | 15 | EPI_ISL_959612 | B.1.1.7 |
| | UKBA-1216/2021 | swab | Belá nad Cirochou | 13.69 | 16 | EPI_ISL_959613 | B.1.1.7 |
| | UKBA-1217/2021 | swab | Bratislava | 14.57 | 17 | EPI_ISL_959614 | B.1.1.7 |
| | UKBA-1218/2021 | swab | Bratislava | 22.37 | 18 | EPI_ISL_959615 | B.1.1.7 |
| | UKBA-1219/2021 | swab | Topoľčany | 15.54 | 19 | EPI_ISL_959616 | B.1.1.7 |
| | UKBA-1221/2021 | swab | Bratislava | 17.71 | 21 | EPI_ISL_959617 | B.1.1.7 |
| | UKBA-1222/2021 | swab | Bratislava | 14.06 | 22 | EPI_ISL_959618 | B.1.1.7 |
| | UKBA-1223/2021 | swab | Bratislava | 24.14 | 23 | EPI_ISL_959619 | B.1.258 |
| | UKBA-1224/2021 | swab | Bratislava | 17.61 | 24 | EPI_ISL_959620 | B.1.258 |
| UKBA-19 | UKBA-1901/2021 | swab | Malacky | 19.45 | 01 | EPI_ISL_1299292 | B.1.1.7 |
| | UKBA-1902/2021 | swab | Malacky | 23.44 | 02 | EPI_ISL_1299293 | B.1.1.7 |
| | UKBA-1903/2021 | swab | Malacky | 23.52 | 03 | EPI_ISL_1299294 | B.1.1.7 |
| | UKBA-1904/2021 | swab | Bratislava | 18.47 | 04 | EPI_ISL_1299295 | B.1.1.7 |
| | UKBA-1905/2021 | swab | Bratislava | 16.57 | 05 | EPI_ISL_1299296 | B.1.1.7 |
| | UKBA-1906/2021 | swab | Bratislava | 21.61 | 06 | EPI_ISL_1299297 | B.1.1.7 |
| | UKBA-1907/2021 | swab | Bratislava | 21.51 | 07 | EPI_ISL_1299298 | B.1.1.7 |
| | UKBA-1908/2021 | swab | Bratislava | 18.15 | 08 | EPI_ISL_1299299 | B.1.1.7 |
| | UKBA-1909/2021 | swab | Bratislava | 22.39 | 09 | EPI_ISL_1299300 | B.1.1.7 |
| | UKBA-1910/2021 | swab | Bratislava | 22.55 | 10 | EPI_ISL_1299301 | B.1.1.7 |
| | UKBA-1911/2021 | swab | Bratislava | 14.16 | 11 | EPI_ISL_1299302 | B.1.1.7 |
| | UKBA-1912/2021 | swab | Bratislava | 24.10 | 12 | EPI_ISL_1299303 | B.1.1.7 |
| | UKBA-1913/2021 | swab | Bratislava | 15.70 | 13 | EPI_ISL_1299304 | B.1.1.7 |
| | UKBA-1914/2021 | swab | Bratislava | 19.90 | 14 | EPI_ISL_1299305 | B.1.1.7 |
| | UKBA-1915/2021 | swab | Malacky | 22.50 | 15 | EPI_ISL_1299306 | B.1.1.7 |
| | UKBA-1916/2021 | swab | Malacky | 25.60 | 16 | EPI_ISL_1299307 | B.1.1.7 |
| | UKBA-1917/2021 | swab | Malacky | 18.60 | 17 | EPI_ISL_1299308 | B.1.1.7 |
| | UKBA-1918/2021 | swab | Malacky | 22.60 | 18 | EPI_ISL_1299309 | B.1.1.7 |
| | UKBA-1919/2021 | swab | Bratislava | 19.60 | 19 | EPI_ISL_1299310 | B.1.1.7 |
| | UKBA-1920/2021 | swab | Bratislava | 23.60 | 20 | EPI_ISL_1299311 | B.1.1.7 |
| | UKBA-1921/2021 | swab | Bratislava | 20.10 | 21 | EPI_ISL_1299312 | B.1.1.7 |
| | UKBA-1922/2021 | swab | Bratislava | 24.40 | 22 | EPI_ISL_1299313 | B.1.1.7 |
| | UKBA-1923/2021 | gargle | Kostoliste | 24.32 | 23 | EPI_ISL_1299314 | B.1.1.7 |
| | UKBA-1924/2021 | gargle | Bratislava | 17.62 | 24 | EPI_ISL_1299315 | B.1.1.7 |
| UKBA-21 | UKBA-2101/2021 | swab | Bratislava | 19.59 | 01 | EPI_ISL_1347634 | B.1.1.7 |
| | UKBA-2102/2021 | swab | Bratislava | 19.25 | 02 | EPI_ISL_1347635 | B.1.1.7 |
| | UKBA-2103/2021 | swab | Malacky | 19.34 | 03 | EPI_ISL_1347636 | B.1.1.7 |
| | UKBA-2104/2021 | swab | Malacky | 19.93 | 04 | EPI_ISL_1347637 | B.1.1.7 |
| | UKBA-2105/2021 | swab | Malacky | 20.14 | 05 | EPI_ISL_1347638 | B.1.1.7 |
| | UKBA-2106/2021 | swab | Malacky | 19.28 | 06 | EPI_ISL_1347639 | B.1.1.7 |
| | UKBA-2107/2021 | swab | Malacky | 18.36 | 07 | EPI_ISL_1347640 | B.1.1.7 |
| | UKBA-2108/2021 | swab | Malacky | 15.65 | 08 | EPI_ISL_1347641 | B.1.1.7 |
| | UKBA-2109/2021 | swab | Malacky | 19.96 | 09 | EPI_ISL_1347642 | B.1.1.7 |
| | UKBA-2110/2021 | swab | Malacky | 18.44 | 10 | EPI_ISL_1347643 | B.1.1.7 |
| | UKBA-2111/2021 | swab | Bratislava | 19.01 | 11 | EPI_ISL_1347644 | B.1.1.7 |
| | UKBA-2112/2021 | swab | Bratislava | 22.94 | 12 | EPI_ISL_1347645 | B.1.1.7 |
| sequencing of three samples shown in grey failed | | | | | | | |

### S1 Fig

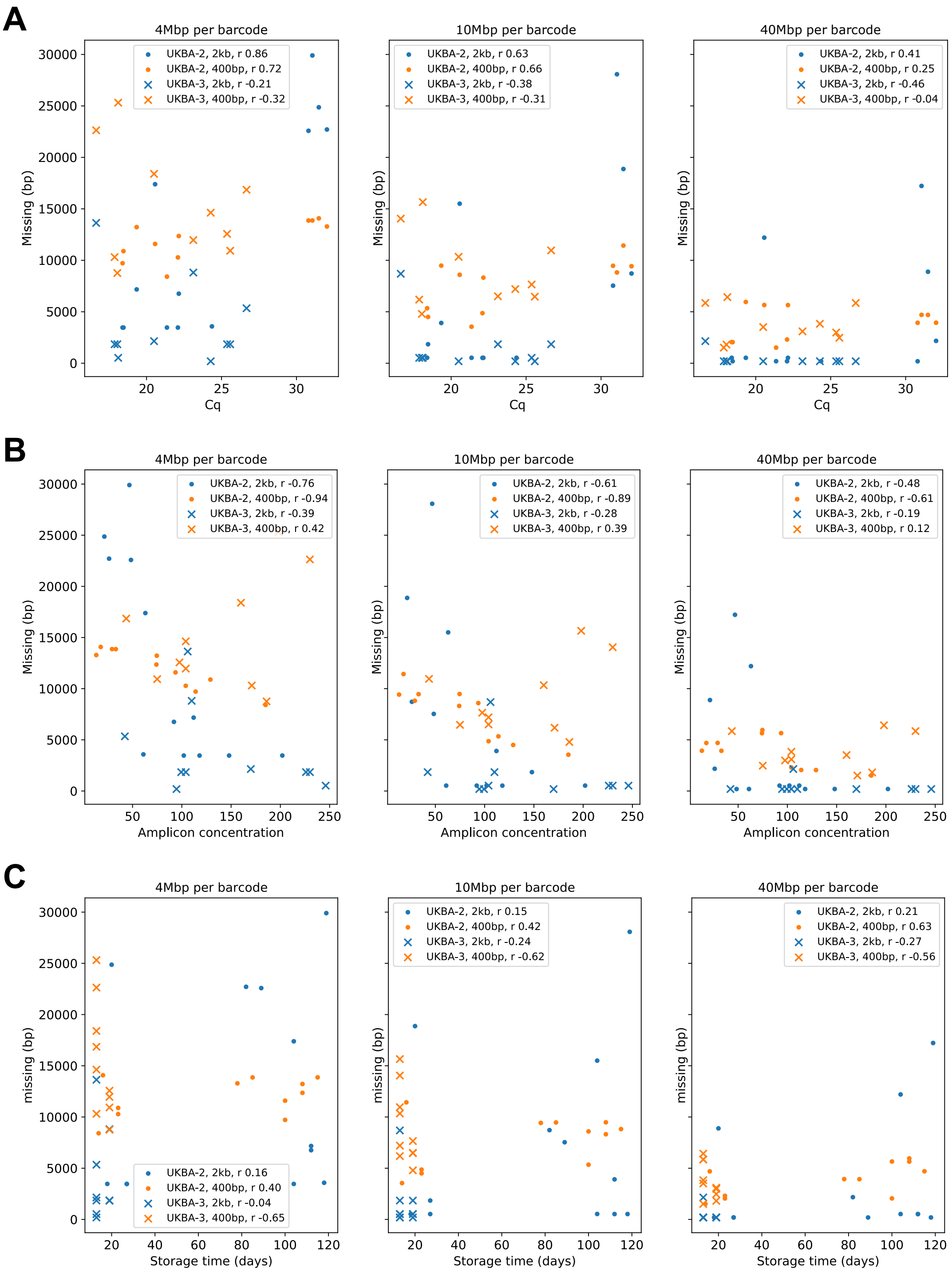
A
4Mbp per barcode
UKBA-2, 2kb, r 0.86
UKBA-2, 400bp, r 0.72
UKBA-3, 2kb, r -0.21
UKBA-3, 400bp, r -0.32
Missing (bp)
Cq
10Mbp per barcode
UKBA-2, 2kb, r 0.63
UKBA-2, 400bp, r 0.66
UKBA-3, 2kb, r -0.38
UKBA-3, 400bp, r -0.31
40Mbp per barcode
UKBA-2, 2kb, r 0.41
UKBA-2, 400bp, r 0.25
UKBA-3, 2kb, r -0.46
UKBA-3, 400bp, r -0.04
B
4Mbp per barcode
UKBA-2, 2kb, r -0.76
UKBA-2, 400bp, r -0.94
UKBA-3, 2kb, r -0.39
UKBA-3, 400bp, r 0.42
Amplicon concentration
10Mbp per barcode
UKBA-2, 2kb, r -0.61
UKBA-2, 400bp, r -0.89
UKBA-3, 2kb, r -0.28
UKBA-3, 400bp, r 0.39
40Mbp per barcode
UKBA-2, 2kb, r -0.48
UKBA-2, 400bp, r -0.61
UKBA-3, 2kb, r -0.19
UKBA-3, 400bp, r 0.12
C
4Mbp per barcode
UKBA-2, 2kb, r 0.16
UKBA-2, 400bp, r 0.40
UKBA-3, 2kb, r -0.04
UKBA-3, 400bp, r -0.65
missing (bp)
Storage time (days)
10Mbp per barcode
UKBA-2, 2kb, r 0.15
UKBA-2, 400bp, r 0.42
UKBA-3, 2kb, r -0.24
UKBA-3, 400bp, r -0.62
40Mbp per barcode
UKBA-2, 2kb, r 0.21
UKBA-2, 400bp, r 0.63
UKBA-3, 2kb, r -0.27
UKBA-3, 400bp, r -0.56

#### S2 Fig

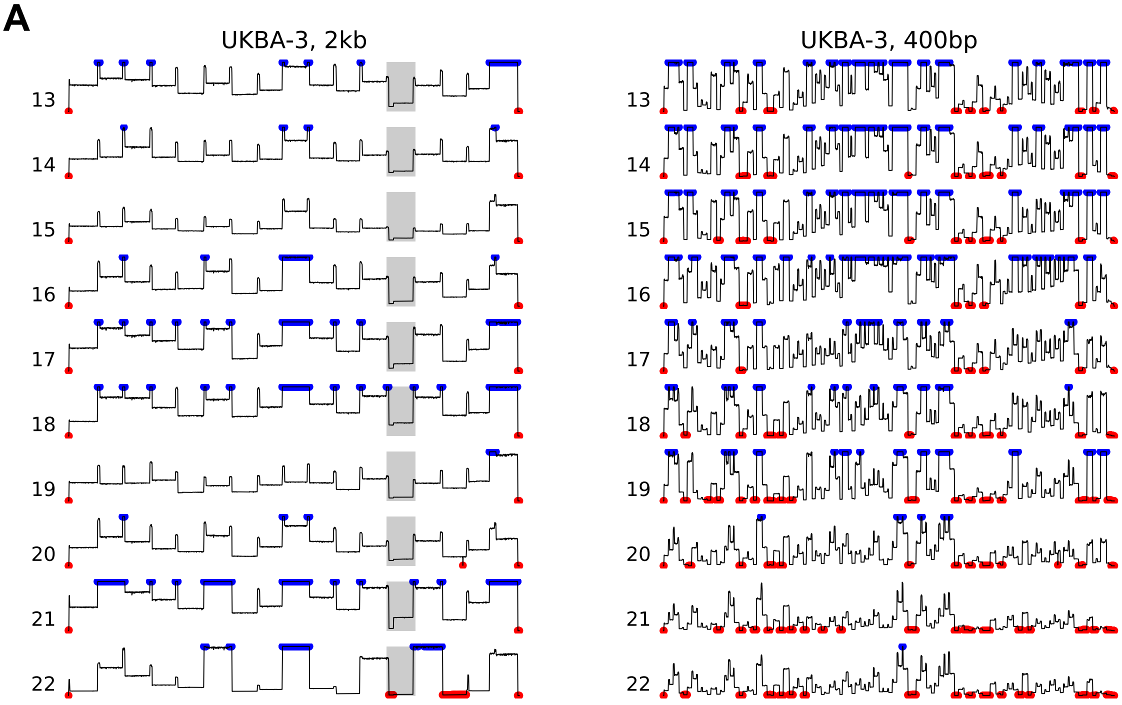
A
UKBA-3, 2kb
UKBA-3, 400bp

**S2 Fig** *(continued)*

**B**

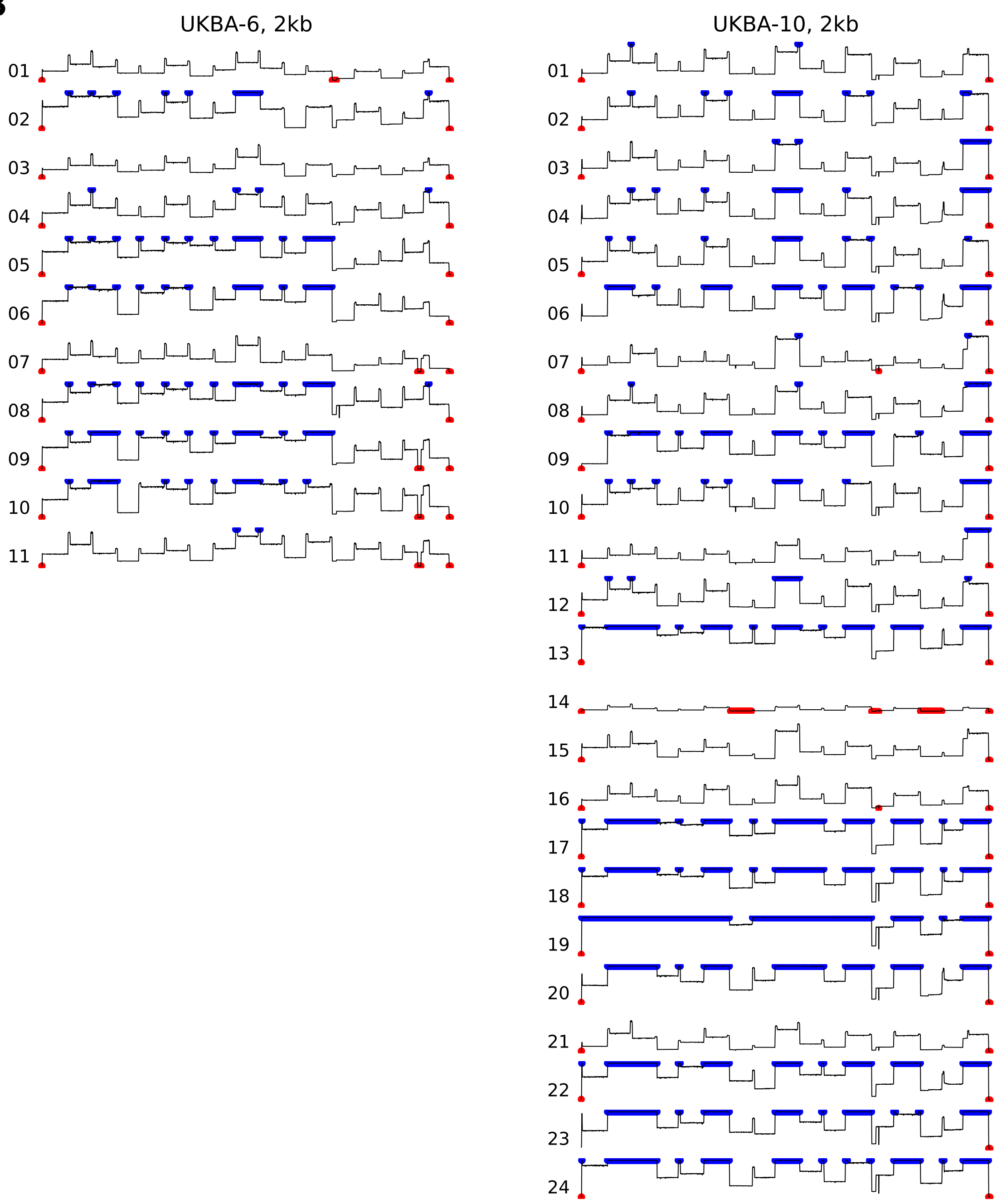

**S2 Fig** *(continued)*

**C**

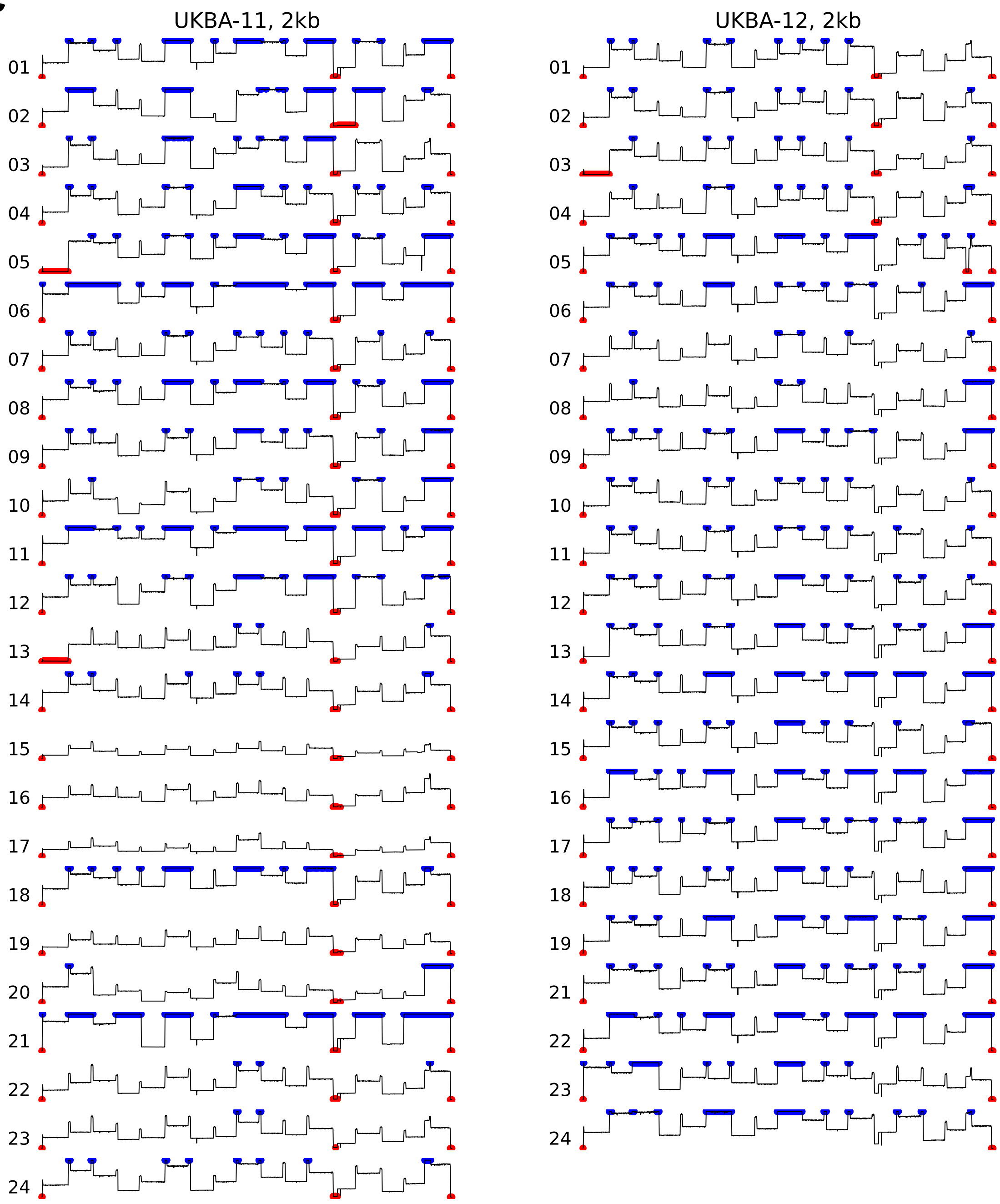

**S2 Fig** *(continued)*

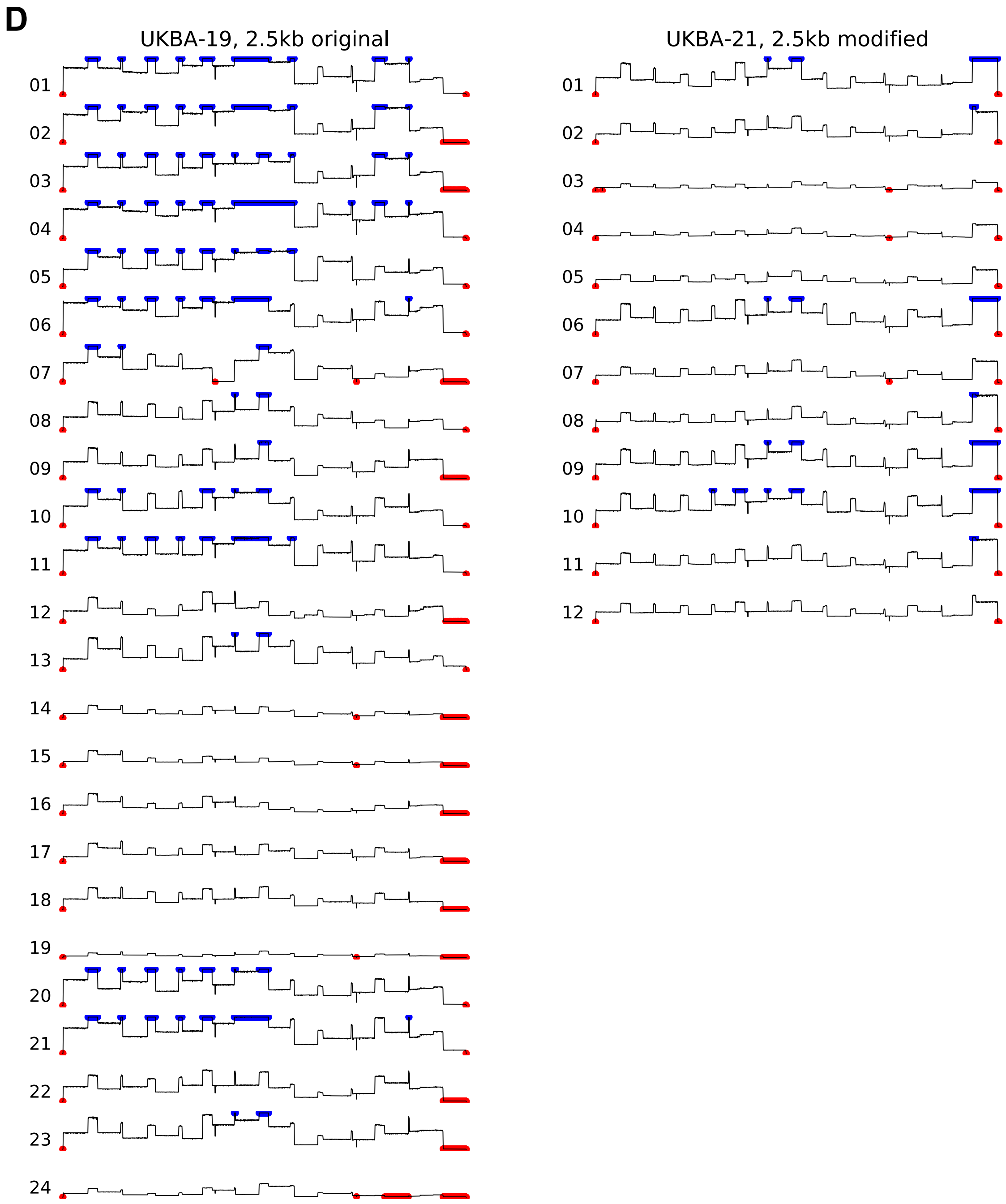
